# Association between Fine Particulate Matter Exposure and Cerebrospinal Fluid Biomarkers of Alzheimer’s Disease among a Cognitively Healthy Population-based Cohort

**DOI:** 10.1101/2023.06.15.23291452

**Authors:** Emma Casey, Zhenjiang Li, Donghai Liang, Stefanie Ebelt, Allan I. Levey, James J. Lah, Thomas S. Wingo, Anke Hüls

## Abstract

**Background:** Epidemiological evidence suggests air pollution adversely affects cognition and increases the risk of Alzheimer’s disease (AD), but little is known about the biological effects of fine particulate matter (PM_2.5_) on early predictors of future disease risk.

**Objectives:** We investigated the association between 1, 3, and 5-year exposure to ambient and traffic-related PM_2.5_ and cerebrospinal fluid (CSF) biomarkers of AD.

**Methods:** We conducted a cross-sectional analysis using data from 1,113 cognitively healthy adults (aged 45-75 years) from the Emory Healthy Brain Study in Georgia, USA. CSF biomarker concentrations of Aβ_42_, tTau, and pTau, were collected at enrollment (between 2016-2020) and analyzed with the Roche Elecsys system. Annual ambient and traffic-related residential PM_2.5_ concentrations were estimated at a 1km and 250m resolution, respectively, and 3- and 5-year average exposures were computed for each participant based on time of specimen collection. Associations between PM_2.5_ and CSF biomarker concentrations, considering continuous and dichotomous (dichotomized at clinical cut-offs for AD-biomarker positivity) outcomes, were estimated with multiple linear/logistic regression, respectively, controlling for potential confounders (age, gender, race/ethnicity, body mass index, and neighborhood socioeconomic status).

**Results:** Interquartile range (IQR; IQR=0.845) increases in 1-year [β: -0.101; 95%-confidence interval (CI): -0.18, -0.02] and 3-year (β: -0.078; 95%-CI: -0.15, -0.00) ambient fine PM_2.5_ exposures were negatively associated with Aβ_42_ CSF concentrations. Associations between ambient PM_2.5_ and Aβ_42_ were similar for 5-year estimates, but not significant (β: -0.076; 95%-CI: -0.160, 0.005). Dichotomized CSF variables revealed similar and significant associations between ambient PM_2.5_ and Aβ_42_. Associations with traffic-related PM_2.5_ were similar but not significant. PM_2.5_ exposures were not associated with tTau, pTau, tTau/Aβ_42_, or pTau/Aβ_42_ levels at enrollment.

**Conclusion:** In our cross-sectional study, PM_2.5_ exposure was associated with a significant decrease in CSF Aβ_42_ which suggests an accumulation of amyloid plaques in the brain and an increased risk of developing AD.

## Introduction

As life expectancy rises and the U.S. population pyramid continues to age, we are seeing an increase in chronic non-communicable age-related conditions, including Alzheimer’s disease and related dementias (AD/ADRD). With the multifactorial nature of AD pathogenesis, building evidence around preventable environmental exposures may ultimately reduce inequities and improve health outcomes. In this regard, accumulating epidemiological evidence demonstrates an association between exposure to air pollution and the prevalence of AD/ADRD. Most of the evidence to date has focused on links between fine particulate matter (PM_2.5_), a mixture of fine particles in the air, and cognitive function, incident cognitive impairment, or dementia.^1-4^ Incident dementia is often studied using diagnostic codes on insurance billing claims and medical records, facilitating studies with large sample sizes; however, billing data are known to miss true dementia cases^1, 5^ and there are no diagnostic codes for the preclinical stages of dementia. In addition, PM_2.5_ can pass through the lung-gas-blood barrier, the gut-brain axis or directly enter brain tissue via the olfactory nerve to promote oxidative stress and inflammation, processes directly related to the characteristic pathology of AD.^6, 7^ Since PM_2.5_ is a heterogeneous mixture, different sources of exposure often have varying degrees of toxicity and most existing studies have focused on ambient exposure, rather than those merely from traffic-related emissions. Understanding the impact of PM_2.5_, from both ambient and traffic-related sources, on preclinical stages of dementia in older at-risk adults is crucial from a public health perspective, as it will improve the estimation of the burden of disease in association with air pollution by identifying more affected individuals.

Recent systematic reviews highlight the need to expand the scope of current studies on air pollution and dementia risk to include neuropathologically relevant outcomes, such as early indicators of dementia, including AD cerebrospinal fluid (CSF) and plasma-based biomarkers, which can be assessed at the late stage of the preclinical phase.^1, 4, 8^ A recent study found positive associations between PM_2.5_ and amyloid β-protein (Aβ)1-40 from plasma in longitudinal analyses, but no associations were detected between PM_2.5_ and Aβ1-42 or the ratio of Aβ1-42/Aβ1-40.^9^ Blood-based biomarkers of protein pathology are less predictive of brain pathology and have shown insignificant changes in Aβ levels as compared to CSF Aβ biomarkers in AD patients.^10^ In this light, three clinically validated CSF biomarkers: Aβ_42_, total tau (tTau), and phosphorylated tau (pTau) have been noted as valid proxies for neuropathological changes of AD,^8, 11^ and specifically, Aβ_42_ has been linked to the abnormal pathologic state of cerebral Aβ in both animal and human models. Since pathological changes related to AD can begin decades before symptoms appear,^12^ quantifying the relationship between both ambient and traffic-related PM_2.5_ pollution and CSF biomarkers reflective of AD-positive changes will elucidate how exposure influences dementia risk.^13^

So far, two epidemiological studies have reported associations between PM_2.5_ and Aβ_42_ in cognitively healthy individuals whereas no relationships with tTau or pTau have been noted.^14, 15^ However, there are several limitations with these prior studies, including 1) the exposure assessments in Alemany *et al*. (2021) and Li *et al*. (2022) only focused on 1- or 2-year average PM_2.5_ prior to the biomarker assessment; 2) the relatively small sample size (N=156) in Alemany *et al*. (2021),^15^ and 3) the outcome assessment in Li *et al*. (2022)^14^ which relied on the Innotest-AMYLOID(1-42) ELISA assay, an unstandardized manual method, showing a high correlation with the automated Elecsys® method but higher intra- and inter-laboratory variations.^16, 17^

To address these limitations in prior studies and grow our understanding of the impact of air pollution on preclinical dementia risk, here, we characterized the cross-sectional association between long-term ambient and traffic-related PM_2.5_ exposure (1, 3, and 5 years prior to biomarker assessment) and CSF biomarker composition (Aβ_42_, tTau, and pTau, assessed with Elecsys^®^ AD CSF assays) in a dementia-free, aging population, as part of the Emory Healthy Brain Study (EHBS). We also tested for effect modification by several well-known risk factors for AD/ADRD-related outcomes, including *APOE*-ε4 status, the strongest genetic risk factor for AD.

## Methods

### Study Design and Population

The EHBS is a gerontology-based prospective research study focusing on the cognitive health of older adults. The EHBS is nested within the Emory Healthy Aging Study (EHAS) and includes participants from the metro-Atlanta region in Georgia, USA. Our cross-sectional analysis includes data from the baseline visits, which were conducted between 2016 and 2020. The primary aim of the EHBS is to characterize psychological and psychosocial factors associated with normal and abnormal aging through assessment of the central nervous system among adults 45-75 years old who were free of cognitive impairment in addition to several other chronic conditions (e.g. congestive heart failure, multiple sclerosis, HIV) at enrollment; more details on recruitment and eligibility have been published elsewhere.^18^

Demographic characteristics were collected with the online Health History Questionnaire (HHQ).^18^ Individual-level information was self-reported for gender, age, race/ethnicity, educational attainment, and residential address. Participants could choose one or more race(s) from a 5-item list (White/Caucasian, Black/African American, Asian, American Indian/Alaska Native, Hawaiian/Other Pacific Islander). Hispanic ethnicity (Yes/No) was addressed in a separate HHQ question. Data on educational attainment were also self-reported with 7 possible categories ranging from less than high school to professional or doctorate degree. EHBS biennial study visits include neuropsychology tests, biospecimen collection (blood, cerebrospinal fluid), cardiovascular measures, and brain imaging. All measures, including anthropometric, are collected by trained clinical research staff for use in the diagnosis and prediction of chronic illness.^18^

### PM_2.5_ Exposure Assessment

Due to evidence suggesting a relationship between both ambient PM_2.5_ and traffic-related PM_2.5_ exposure and cognitive decline, and because traffic-related PM_2.5_ is a major exposure source in urban environments like Atlanta, GA,^19^ we used both measures of PM_2.5_ in our analyses.

We obtained ambient PM_2.5_ exposure data from the publicly available Socioeconomic Data and Application Center (SEDAC) air quality data set for health-related applications.^20^ The data set consists of yearly ambient PM_2.5_ levels (in μg/m^3^) estimated at a 1 km spatial resolution using a well-validated ensemble-based prediction model for the contiguous United States (2000-2016). As described by Di *et al*. (2019), three machine-learning algorithms: random forest, neural network, and gradient boosting, were used to predict ambient PM_2.5_ and included a variety of predictor variables from satellite data, land use, meteorological variables, and chemical transport model simulations. ^21^

The ensemble model then combined these PM_2.5_ predictions with a generalized additive model that allowed for the contribution of each machine-learning algorithm to vary by location.^21^ The ensemble model was trained on PM_2.5_ levels measured at 2,156 U.S. EPA monitors, validated with 10-fold cross-validation, and produced high-resolution annual PM_2.5_ predictions with an average R^2^ of 0.89.^21^

As described previously,^22^ traffic-related PM_2.5_ exposure concentrations (in μg/m^3^) for the metro-Atlanta area for the period 2002 to 2019 were generated by two approaches. Briefly, for the 2002-2011 period, annual traffic-related PM_2.5_ concentrations at 250 m resolution were generated by the Research LINE-sources dispersion (R-LINE) model and calibrated by measurements from local PM_2.5_ ground monitoring sites.^23, 24^ For the 2012-2019 period, annual traffic-related PM_2.5_ concentrations at 200-250 m resolution were predicted via a land-use random forest model built on training data comprised of the 2015 annual concentrations of traffic-related PM_2.5_ from Atlanta Reginal Commission, road inventory and traffic monitoring data based on measurements from the Georgia Department of Transportation which considered road geometry and traffic volume, land cover data from the National Land Cover Database, and ambient PM_2.5_ data from Atmospheric Composition Analysis. The random forest model was trained with the R package *randomForest*. The resulting 200-250m resolution annual traffic-related PM_2.5_ predictions had an average R^2^ of 0.80.

Both databases had spatial coverage of approximately 20 counties in GA. Given the reduced coverage of the traffic-related PM_2.5_ exposure estimates compared to the ambient PM_2.5_ data and the geographic spread of our study population, the analysis sample for these models included only the participants located within the metro-Atlanta area.

For both ambient and traffic-related exposures, we spatially matched geocoded residential addresses to the closest centroid of grids (based on 1 km^2^ or 200-250 m^2^ grids) to assign annual exposures. We then calculated individual 3- and 5-year exposures by averaging yearly predictions prior to specimen collection.

### AD CSF Biomarker Concentrations

CSF biospecimens were collected by EHBS research staff via lumbar puncture, CSF collection protocol has been previously described.^25^ Aβ_42_, tTau, and pTau CSF levels were quantified using the ElectroChemiLuminescense Immunoassay (ECLIA) Elecsys® AD CSF portfolio on an automated Roche Diagnostics instrument (F. Hoffman-La Roche Ltd). The assays have measuring ranges of 200–1700 pg/mL (Aβ_42_), 80–1300 pg/mL (tTau) and 8–120 pg/mL (pTau). tTau and pTau levels were log-base_10_ transformed for normality in linear models in the statistical analyses. We also examined CSF biomarker ratio outcomes, namely, tTau/Aβ_42_ and pTau/Aβ_42_, which are highly predictive of amyloid positivity based on concordance with amyloid-PET, including for cognitively normal participants.^26^ All AD CSF biomarker outcomes were kept as continuous variables for linear regression analyses and separately dichotomized based on the Elecsys® AD CSF portfolio positive (+) cut-offs for logistic regression analyses (Aβ_42_ ≤1030 pg/mL; tTau >300 pg/mL; pTau >27 pg/mL; tTau/Aβ_42_ >0.28pg/mL; pTau/Aβ_42_ >0.023pg/mL).^8, 27^ The cut-off values for Aβ_42_, tTau/Aβ_42_, and pTau/Aβ_42_ CSF were established and validated to demonstrate CSF biomarker concordance with amyloid-β PET visual read in the BioFINDER and ADNI studies, respectively.^28^ Cut-off values for single tau biomarkers, tTau and pTau, were derived and validated in a separate study based on the separation between mild cognitive impairment (MCI) patients with a higher versus lower risk of cognitive decline, and optimized for identification of AD patients versus normal controls.^29^

### Covariates

Sources of potential confounding were identified with a directed acyclic graph (DAG; **Figure S1**). Individual-level confounders were conceptualized as factors impacting both residential PM_2.5_ exposure and the outcome measure. Potential confounding factors included in the analysis were gender, age, neighborhood socioeconomic status (N-SES), race/ethnicity, educational attainment, and body mass index (BMI). Due to historic racism and discriminatory land-use practices such as redlining, environmental exposures disproportionately affect low-income and minority populations. For this reason, neighborhood deprivation characteristics were also included as potential confounding variables and effect modifiers as done in our previous work.^30^ Race has also been noted as an important factor when interpreting CSF biomarker results.^31^ In addition, BMI influences biomarker concentrations and is also related to N-SES through characteristics such as neighborhood walkability, greenspace, and food access.^32, 33^

Due to the presence of multi-ancestral groups and small categories in self-reported race/ethnicity, we used a 3-level race variable in the analysis: White/Caucasian, Black/African American, and Other, as well as a dichotomous ethnicity variable indicating Hispanic origin. Similarly, educational attainment was included as a 3-level variable: master or higher, college, less than college. Height and weight measurements were used to calculate body mass index (BMI, weight in kilograms divided by height in meters squared) which was used as a continuous variable in all models. N-SES for each participant was established in this study with census-tract level American Community Survey (ACS) defined principal components of neighborhood deprivation (see Li *et al*.^30^ for details) and the Area Deprivation Index (ADI). As described previously in Li *et al*.,^30^ three principal components of neighborhood deprivation were calculated based on estimates for 5-year ACS census-tract-level data, including 16 indicators of six socioeconomic domains (poverty/income, racial composition, education, employment, occupation, and housing properties) (**Figure S2**).^30^ The ADI is provided in national percentile rankings at the block group level from 1 to 100, where 100 represents the most deprived neighborhood, and was calculated using census block group-level indicators and factor analysis to cluster indicators based on their ability to explain the variance between block groups.^34^

### Statistical Analyses

We implemented multiple linear and logistic regression models to estimate the relationship between residential PM_2.5_ exposure and AD CSF biomarker levels at enrollment. In these models, biomarker concentrations (linear regression models) or dichotomized AD positive variables (logistic regression models) were assigned as dependent variables and PM_2.5_ exposures along with the selected confounding variables as independent variables. Since the biomarkers were measured on the same scales, but with different ranges, we standardized all continuous biomarker measures by converting them to z-scores prior to employing regression analysis to increase comparability of results across different biomarkers. Z-scores were computed for each observation by subtracting the sample mean from each individual value and subsequently dividing by the sample standard deviation. Further, we standardized the PM_2.5_ estimates according to its distribution by dividing all PM_2.5_ exposures of interest by the IQR of 1-year ambient or traffic-related PM_2.5_ exposure respectively. The general form of the model for all analyses appears below:

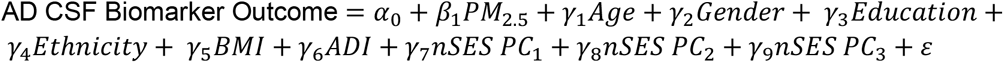

Where PM_2.5_ represents either ambient or traffic-related PM_2.5_ averages 1, 3, or 5 years prior to specimen collection and ε represents the random error term, with an assumed mean of zero and constant variance ∼N(0, σ^2^).

### Effect Modification Analyses

We tested for effect modification by several well-established risk factors for AD, adding an interaction term between PM_2.5_ and each risk factor in individual regression models. These risk factors included whether a participant had at least one *APOE*-ε4 allele, family history of AD (indicated by parent or first-sibling diagnosis), gender, age, and ADI. Despite a hypothesized additive genetic effect of the ε4 allele, statistical interaction was assessed dichotomously due to the small number of homozygous ε4 carriers (2.1%; **Table 1**). Family history of AD (no/yes), gender (male/female), and ADI (<50/≥50) were also added as dichotomous variables while interaction with age was assessed continuously. Using the models with interaction, we then tested for effect modification with the *interplot* R package and n=100,000 simulations.

**Table 1.** Baseline descriptive characteristics for EHBS study participants (≥45 years of age).

| Characteristics<sup>d</sup> | Overall<br>(N=1113) |
| --- | --- |
| **Age (years)** |  |
| Mean (SD) | 61.7 (6.73) |
| Median [Min, Max] | 62.0 [45.0, 77.0] |
| **Gender** |  |
| Female | 775 (69.6%) |
| **Body Mass Index (kg/m<sup>2</sup>)** |  |
| Mean (SD) | 25.5 (3.55) |
| Median [Min, Max] | 25.3 [16.8, 38.4] |
| **Race/Ethnicity<sup>a</sup>** |  |
| White/Caucasian | 943 (84.7%) |
| Black/African American | 121 (10.9%) |
| Other | 49 (4.4%) |
| **Hispanic** |  |
| Yes | 32 (2.9%) |
| **Educational Attainment<sup>b</sup>** |  |
| Less than college | 147 (13.2%) |
| College | 478 (42.9%) |
| Master or higher | 488 (43.8%) |
| **Area Deprivation Index<sup>c</sup>** |  |
| Mean (SD) | 29.4 (20.1) |
| Median [Min, Max] | 25.0 [1.00, 93.0] |
| **APOE-ε4 allele carriership** |  |
| No allele | 591 (53.1%) |
| 1 allele | 241 (21.7%) |
| 2 alleles | 23 (2.1%) |
| Missing | 258 (23.2%) |
| **<u>Air pollution concentration</u>** |  |
| **1 yr. ambient PM<sub>2.5</sub> (μg/m<sup>3</sup>)** |  |
| Mean (SD) | 9.52 (0.764) |
| Median [Min, Max] | 9.52 [5.63, 13.2] |
| IQR | 0.845 |
| **3 yr. ambient PM<sub>2.5</sub> (μg/m<sup>3</sup>)** |  |
| Mean (SD) | 9.85 (0.832) |
| Median [Min, Max] | 9.96 [5.99, 12.0] |
| IQR | 1.10 |
| **5 yr. ambient PM<sub>2.5</sub> (µg/m<sup>3</sup>)** |  |
| Mean (SD) | 9.96 (0.735) |
| Median [Min, Max] | 10.1 [6.41, 12.0] |
| IQR | 0.945 |
| **1 yr. traffic PM<sub>2.5</sub> (µg/m<sup>3</sup>)** |  |
| Mean (SD) | 1.15 (0.464) |
| Median [Min, Max] | 1.10 [0.155, 5.06] |
| IQR | 0.523 |
| Missing *n* (%) | 33 (3.0%) |
| **3 yr. traffic PM<sub>2.5</sub> (µg/m<sup>3</sup>)** |  |
| Mean (SD) | 1.26 (0.455) |
| Median [Min, Max] | 1.23 [0.175, 5.56] |
| IQR | 0.479 |
| Missing *n* (%) | 33 (3.0%) |
| **5 yr. traffic PM<sub>2.5</sub> (µg/m<sup>3</sup>)** |  |
| Mean (SD) | 1.25 (0.432) |
| Median [Min, Max] | 1.21 [0.168, 5.38] |
| IQR | 0.447 |
| Missing *n* (%) | 33 (3.0%) |

<sup>a</sup>Self-reported race/ethnicity was categorized into 3 groups: White/Caucasian, Black/African American, and Other which includes both minority and mixed ancestry groups.
<sup>b</sup>Educational attainment was self-report and included less than high school, high school/GED, some college, associate's degree, bachelor's degree, master's degree, and a professional degree (e.g., Ph.D.).
<sup>c</sup>Derived from factor analysis and validated to the Census Block Group neighborhood level with factors for the theoretical domains of income, education, employment, and housing quality; higher scores indicate higher levels of "disadvantage."
<sup>d</sup>Abbreviations: yr., Year; SD, standard deviation; Min, minimum; Max, maximum; IQR, interquartile range; µg, Microgram; m<sup>3</sup>, meters cubed.

For all statistical analysis, we used R Statistical Software version 4.2.2, and the significance level α=0.05.

## Results

### Study Population

After excluding EHBS participants with missing demographic data (n=46), the analytic sample included 1,113 individuals (**Table 1)**. Participants lived primarily around the Atlanta metropolitan area, spanning 489 census blocks in the state of Georgia (detailed neighborhood characteristics are summarized in **Table S6**). The average age of our sample was 61.7 years (SD=6.73), 69.9% were females, and 84.7% identified as White/Caucasian, 10.9% identified as Black/African American, and only 2.9% identified as Hispanic. The average BMI was 25.5 kg/m^2^ (SD=3.55). Our sample was highly educated with 86.7% having received an associate degree or higher. The ADI was right skewed with a median of 25, indicating half of the sample is in 25% lower deprivation than the national average.

There was spatial variability in ambient PM_2.5_ levels in our study area, with the highest quintile of exposure (10.1-13.21 μg/m^3^) localized to the south of the city of Atlanta and lowest quintile of exposure (5.63-8.98 μg/m^3^) localized to communities north of Atlanta such as Marietta and Roswell (**Figure 1, Table 1**). Traffic-related PM_2.5_ exposure levels had lower concentrations (since we only estimated the traffic-related component of PM_2.5_) but higher variability with annual average exposures of 1.15 μg/m^3^ (SD=0.46) compared with annual average ambient exposures of 9.52 μg/m^3^ (SD=0.76). Traffic-related PM_2.5_ estimates had a maximum of 5.10 μg/m^3^ and these levels were localized exclusively to the city of Atlanta. Annual ambient and traffic-related PM_2.5_ exposure concentrations were weakly correlated (Pearson correlation = 0.36). More details on the distribution of and relation between ambient and traffic-related PM_2.5_ exposure concentrations are provided in the supplemental material (**Figures S3 and S4**).

**Figure 1.**
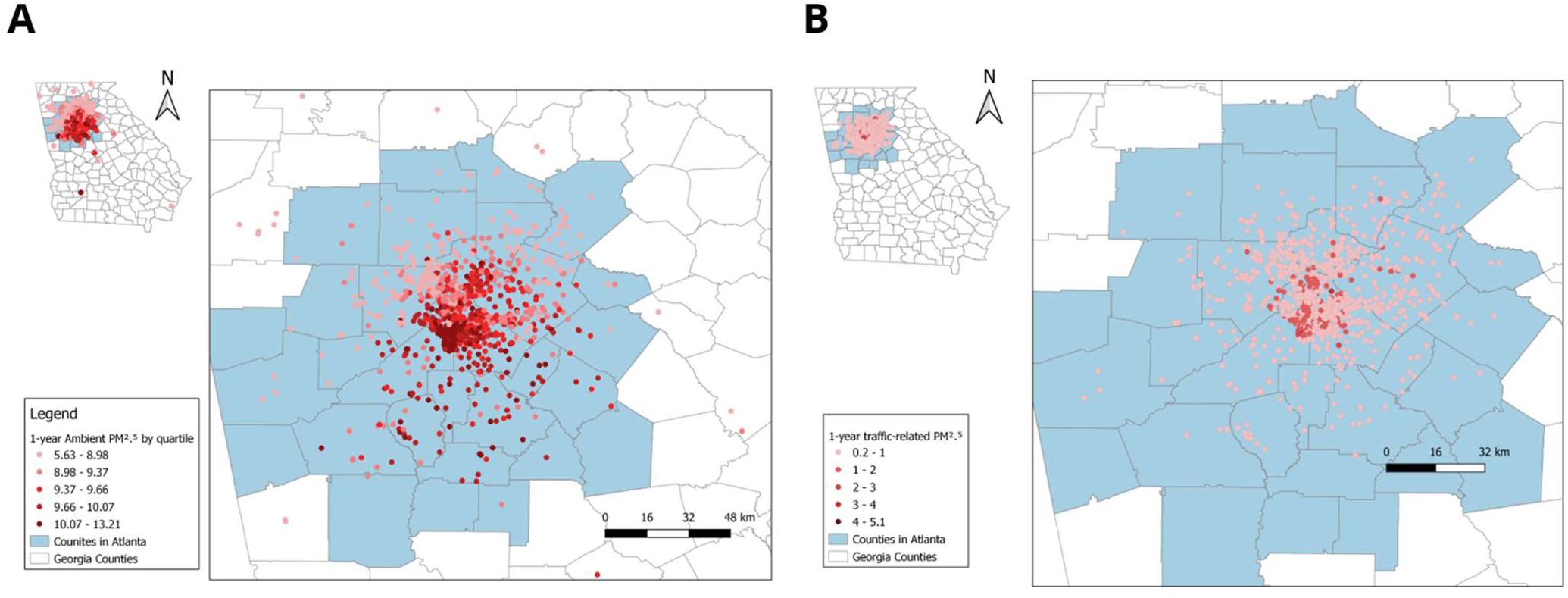
Map of the geographic distribution, each dot representing a study participant, of our study population and annual ambient residential PM_2.5_ (in μg/m^3^) exposure by quantile (**A**) (N=1,113) or annual traffic-related residential PM_2.5_ exposure (min-max) with easy breaks (**B**) (N=1,080).

We observed a wide spread of concentrations for CSF Aβ_42_ in the study population (median Aβ_42_ level = 1210, IQR = 692.3). Aβ_42_ concentrations did not show a major departure from normality, except for the highest level which had a very high frequency, indicating normal Aβ_42_ concentrations in most participants, but tTau and pTau distributions were skewed (**Figure S5**). After log transformation, Tau concentrations were approximately normally distributed (**Figure S6**).

Approximately 36% of participants had Aβ_42_ concentrations less than or equal to 1030 pg/mL which corresponds, on average, to a positive reading for AD as indicated by Elecsys^®^ AD CSF portfolio positive (+) cut-offs. We observed AD positive readings for tTau and pTau cut-offs in 6% of the study population. Based on the pTau/Aβ_42_ ratio AD (+) positive cut-off, we detected amyloid-positivity in 10.6% of participants. Details on the distributions of AD CSF biomarker concentrations and the frequency of biomarker-positivity detected among participants are provided in **Table 2**.

**Table 2.** Baseline AD CSF Outcomes at enrollment between 2016 and 2020.

| AD CSF concentrations and (+) cut-off | Total<br>(N=1113) |
| --- | --- |
| **Aβ<sub>42</sub> (pg/mL)** |  |
| Mean (SD) | 1200 (382) |
| Median [Min, Max] | 1210 [200, 1700] |
| IQR | 692.3 |
| **tTau (pg/mL)** |  |
| Mean (SD) | 187 (70.9) |
| Median [Min, Max] | 174 [80.0, 799] |
| IQR | 79.6 |
| **pTau (pg/mL)** |  |
| Mean (SD) | 16.7 (7.16) |
| Median [Min, Max] | 15.2 [8.00, 83.8] |
| IQR | 7.63 |
| **tTau/Aβ<sub>42</sub> (pg/mL)** |  |
| Mean (SD) | 0.171 (0.107) |
| Median [Min, Max] | 0.141 [0.0818, 1.72] |
| **pTau/Aβ<sub>42</sub> (pg/mL)** |  |
| Mean (SD) | 0.0154 (0.0116) |
| Median [Min, Max] | 0.0123 [0.00692, 0.200] |
| **Aβ<sub>42</sub>** |  |
| (+) | 402 (36.1%) |
| **tTau** |  |
| (+) | 68 (6.1%) |
| **pTau** |  |
| (+) | 65 (5.8%) |
| **tTau/Aβ<sub>42</sub>** |  |
| (+) | 87 (7.8%) |
| **pTau/Aβ<sub>42</sub>** |  |
| (+) | 118 (10.6%) |

<sup>a</sup>AD CSF portfolio positive (+) vs (-) cut-offs (Aβ <sub>42</sub> ≤1030 pg/ml; tTau >300 pg/mL; pTau >27 pg/mL; tTau/Aβ <sub>42</sub> >0.28pg/mL; pTau/Aβ <sub>42</sub> >0.023pg/mL)

### PM_2.5_ and AD CSF biomarkers

Among the continuous AD CSF biomarkers, higher levels of 1- and 3-year ambient PM2.5 exposures were associated with lower Aβ_42_ CSF concentrations at baseline after adjusting for potential confounding variables (**Figure 2A**). Specifically, an IQR (0.845 μg/m^3^) increase in the 1- or 3-year ambient PM_2.5_ exposure was significantly associated with a -0.09 (95% CI: -0.15, -0.02) and -0.07 (95% CI: -0.13, -0.005) decrease in Aβ_42_ CSF z-score, respectively, after confounder-adjustment. The associations of 5-year ambient PM_2.5_ (**Figure 2A**) and traffic-related PM_2.5_ (**Figure 2F**) and Aβ_42_ CSF were similar, but not significant. No significant associations were detected between ambient (**Figure 2B-E**) or traffic-related (**Figure 2G-J**) PM_2.5_ exposures and tTau, pTau, tTau/Aβ_42_, or pTau/Aβ_42_ CSF concentrations at enrollment, but associations with tTau/Aβ_42_ and pTau/Aβ_42_ were similar to those of Aβ_42_ and consistent with AD-related pathology (**Figure 2I-J**).

**Figure 2.**
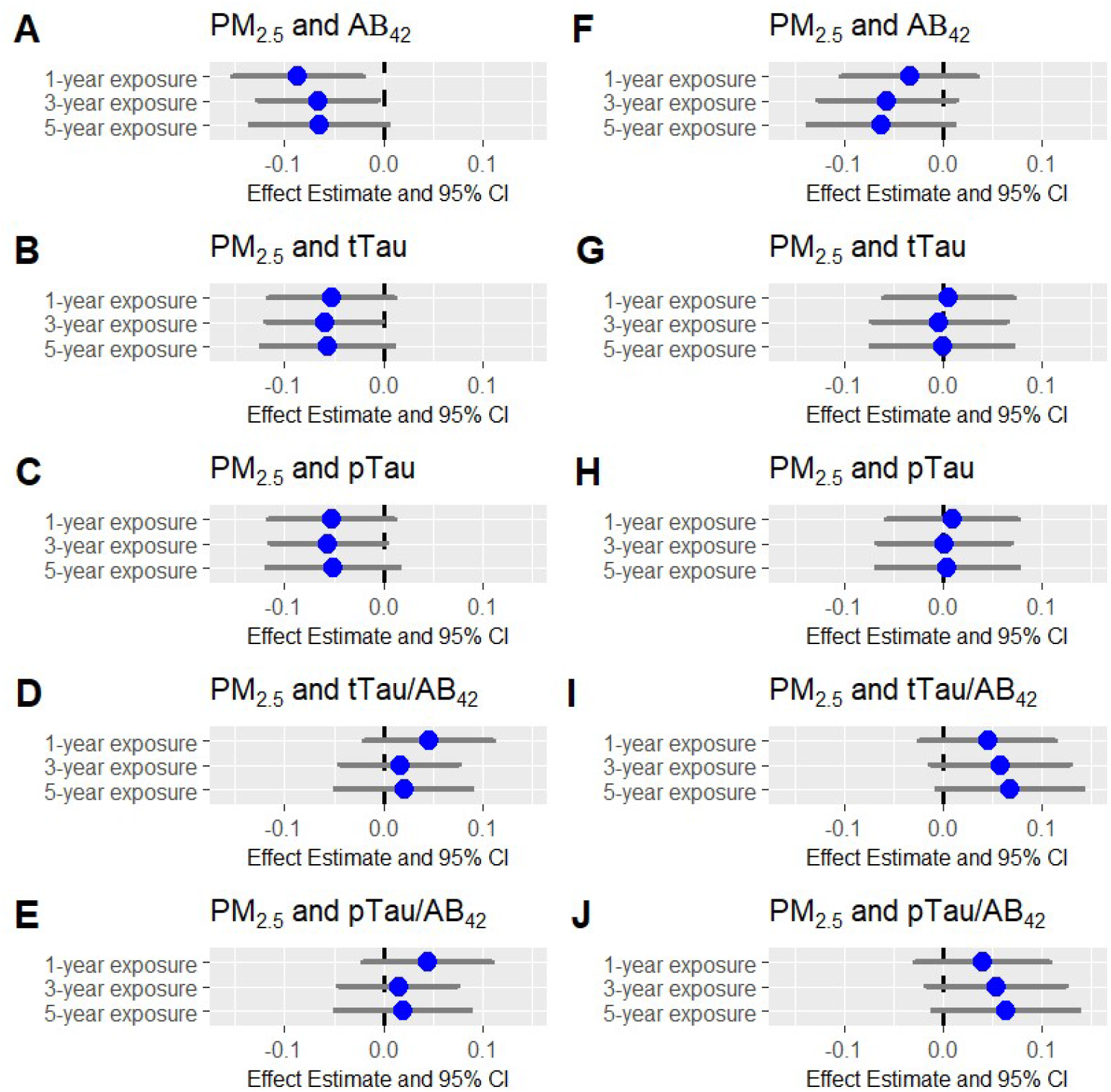
Effect Estimate (± 95% CI) of 1, 3, and 5-year ambient (**A**–**E**) (N=1,113) and traffic-related (**F**–**J**) (N=1,080) PM_2.5_ exposure on AD CSF Biomarker concentrations (in pg/mL) (Aβ_42_, tTau, pTau, tTau/Aβ_42_, and pTau/Aβ_42_). All estimates are standardized and adjusted for gender, age, N-SES, ethnicity, educational attainment, and BMI. The dashed line indicates the significance threshold: 0 for linear regression.

For the AD CSF biomarker positive cut-off outcomes, higher residential ambient PM_2.5_ exposures were associated with increased prevalence of an AD positive (+) Aβ_42_ portfolio reading at baseline with statistically significant associations for an IQR (0.845 μg/m^3^) increase in 1-year (OR=1.23; 95% CI: 1.07, 1.42), 3-year (OR=1.20; 95% CI: 1.04, 1.37), and 5-year (OR=1.21; 95% CI: 1.04, 1.40) average ambient PM_2.5_ exposure (**Figure 3A**). The associations between traffic-related PM_2.5_ exposures and an AD positive (+) Aβ_42_ portfolio reading at enrollment were similar but not significant (**Figure 3F**). We further observed significant associations between 3- and 5-year traffic-related PM_2.5_ exposures and an AD (+) pTau/Aβ_42_ ratio reading at enrollment, but not for 1-year traffic-related PM_2.5_ exposure (**Figure 3J**) or ambient PM_2.5_ exposure (**Figure 3E**), and while not significant, these results were consistent with those of tTau/Aβ_42_ (**Figure 3I, Figure 3D**). We did not observe statistically significant associations between ambient (**Figure 3B-C**) or traffic-related (**Figure 3G-H**) PM_2.5_ exposures and AD positive (+) tTau or pTau cut-offs.

**Figure 3.**
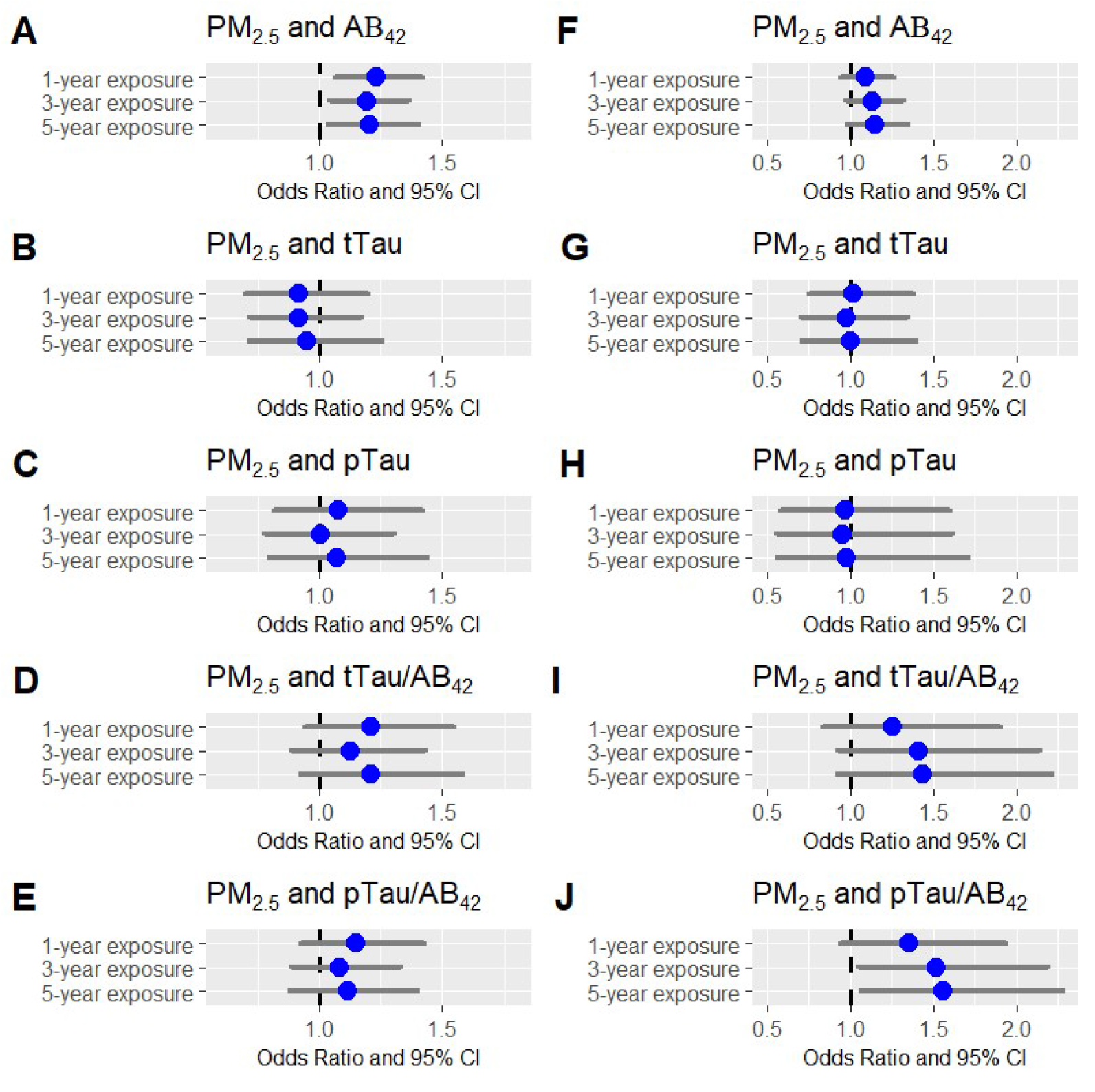
Odds ratio (± 95% CI) for the association between 1, 3, and 5-year ambient (**A**–**E**) (N=1,113) and traffic-related (**F**–**J**) (N=1,080) PM_2.5_ exposure and AD CSF Biomarkers positive (+) cut-offs (Aβ_42_, tTau, pTau, tTau/Aβ_42_, and pTau/Aβ_42_). All estimates are adjusted for gender, age, N-SES, ethnicity, educational attainment, and BMI. The dashed line indicates the significance threshold: 1 for logistic regression.

The associations between ambient PM_2.5_ and Aβ_42_ CSF concentrations and AD positive (+) Aβ_42_ portfolio reading remained significant even after restricting our sample size to only those located in the metro-Atlanta area (**Figure S7, Tables S1, S2**, and **S3**), which is the subsample for which traffic-related PM_2.5_ exposures estimates were available (sample size reduced from N_ambient_=1,113 to N_traffic_=1,080).

### Effect modification by other common risk factors for AD

The association of annual average ambient PM_2.5_ exposure and concentrations of Aβ_42_ CSF was not significantly modified by *APOE*-ε4 carriership (p = 0.59), AD family history (p = 0.37), ADI (p = 0.62) or gender (p = 0.67) (**Figure 4A-D**, **Table S4**). Similarly, the association of annual average ambient PM_2.5_ exposure and AD positive (+) Aβ_42_ portfolio reading was not significantly modified by *APOE*-ε4 carriership (p = 0.80), AD family history (p = 0.27), ADI (p = 0.12) or gender (p = 0.72) (**Figure S8A- D**, **Table S5**). Effect modification by age was also not statistically significant (p=0.17), but we observed an increasing negative effect of PM_2.5_ on Aβ_42_ CSF levels with increasing age, and significant associations between PM_2.5_ and Aβ_42_ CSF levels starting around 60 years of age (**Figure 4E**); a similar pattern was revealed when looking at the stratified effects of PM_2.5_ on AD positive (+) Aβ_42_ portfolio reading by age (interaction p=0.34) (**Figure S8E**).

**Figure 4.**
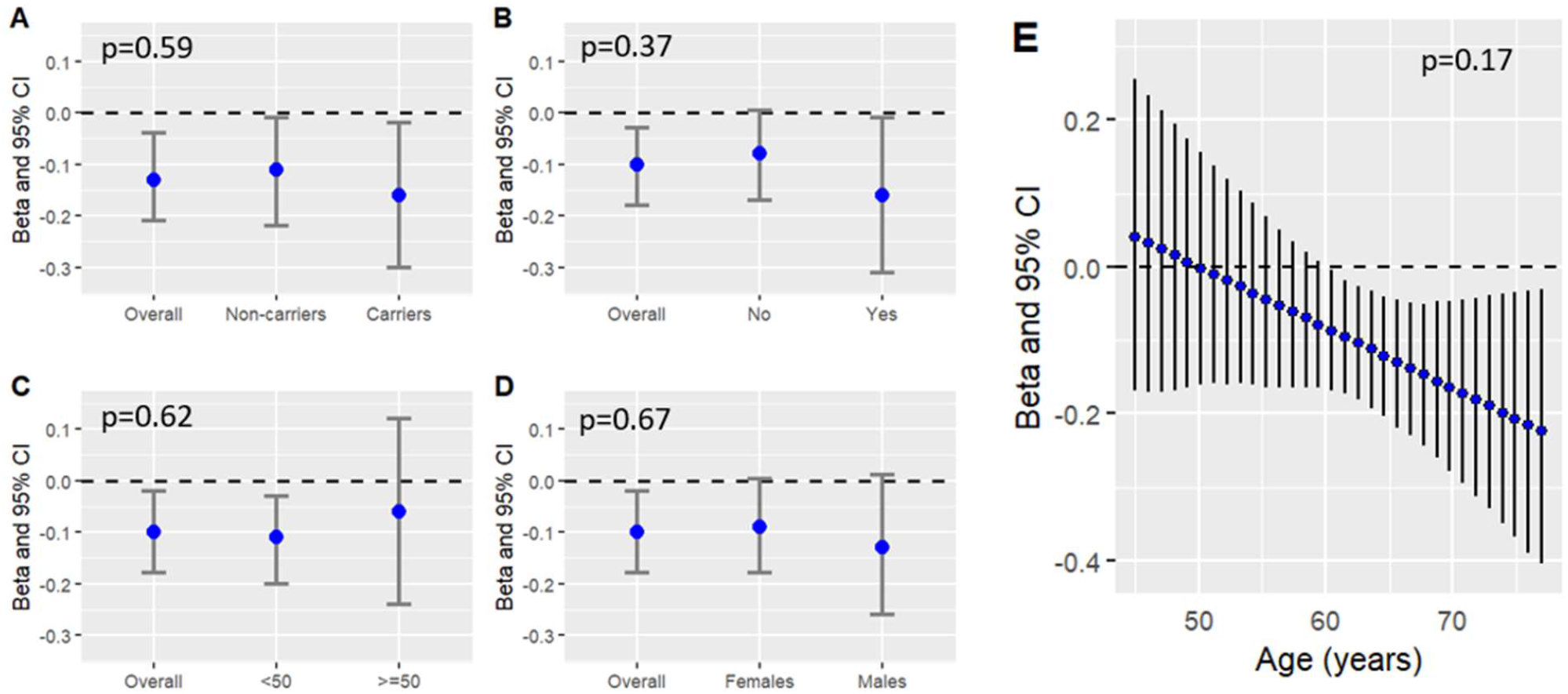
Effect (± 95% CI) of yearly ambient PM_2.5_ exposure on Aβ_42_ CSF concentrations (in pg/mL) by *APOE*-ε4 carriership (**A**), AD family history (**B**), ADI (**C**), Gender (**D**), and Age (**E**). Presented as overall and stratified effects for dichotomous variables and as continuous for Age, with interaction p-values depicted on each graph. The dashed line indicates the significance threshold: 0 for linear regression. The overall effect in **Figure 4A** (N=855) differs slightly from **Figure 4B-D** (N=1113) due to the decreased sample size after including only participants with *APOE* genotype data.

## Discussion

In the present study, we examined the impacts of both ambient PM_2.5_ exposure and traffic-related PM_2.5_, a major source of ambient PM_2.5_ in urban environments, on CSF biomarkers of AD in 1,113 cognitively healthy individuals. Our findings show associations between long-term ambient PM_2.5_ concentrations and decreased Aβ_42_ AD CSF biomarker concentrations (significant for 1- and 3-year average exposures), as well as increased likelihood of an Aβ_42_ AD (+) positive portfolio reading (significant for 1-, 3- and 5-year average exposures). We further found significant associations between 3- and 5-year traffic-related PM_2.5_ exposure estimates and a pTau/Aβ_42_ (+) positive portfolio reading at enrollment, but not with ambient PM_2.5_ exposure. We found no associations between ambient or traffic-related PM_2.5_ exposures and pTau or tTau continuous concentrations or their ratios with Aβ_42_, however, the directions of effect for pTau/Aβ_42_ and tTau/Aβ_42_ continuous ratio outcomes were consistent with AD-related amyloid pathology. Further, while not statistically significant, the strength of the association between annual ambient PM_2.5_ exposure and Aβ_42_ AD CSF concentrations differed by age and was particularly pronounced for individuals over the age of 60.

The observed associations between PM_2.5_ exposure and the Aβ_42_ AD CSF biomarker as well as pTau/Aβ_42_ positive portfolio readings, which are equally predictive of amyloid PET status (+/-) as Aβ ratio outcomes,^35^ among cognitively healthy older adults is consistent with evidence from existing literature. Signs of AD can be detected in the early stages of the AD continuum,^17^ and decreases in CSF concentrations of Aβ_42_ (a marker of amyloidosis) and elevation in tau species (phosphorylated and total tau) are well-established as pathogenic biomarkers in AD diagnosis.^36^ To date, there have been few studies estimating the effects of PM_2.5_ exposure on certified biomarkers of AD in healthy, aging populations. One study found a similar relationship between air pollution exposure and Aβ_42_, although they used CSF Aβ_42/40_ ratio to reflect Aβ pathology rather than the individual biomarker measurements.^15^ Their estimates were similarly negative, but did not reach significance, likely owing to the relatively small sample size (N=147).^15^ Another study^14^ found a statistically significant total effect of ambient PM_2.5_ on Aβ_42_ CSF as well as pTau/Aβ_42_ concentrations among N=1,131 cognitively healthy older individuals, which was further mediated by a CSF biomarker of neuroinflammation, sTREM2.^14^

We found mostly null associations between 1-, 3-, and 5-year average ambient and traffic-related PM_2.5_ exposures and tTau as well as pTau concentrations at enrollment, except for the model with 3-year ambient PM_2.5_ and tTau levels, in which we found a significant negative association. The direction of estimates for tau biomarkers are expected to be positive for AD-related change.

Associations between PM_2.5_ and tau biomarkers diminished when taking AD CSF cut-offs into account, suggesting that the negative association with the continuous tTau levels was a false positive finding. Other studies examining the associations between air pollution and CSF biomarkers of AD in cognitively healthy adults report similar findings, where both Alemany *et al*. (2021) and Li *et al*. (2022) found null associations between PM_2.5_ and pTau as well as tTau CSF concentrations.^14, 15^

Stronger associations were detected between ambient PM_2.5_ and AD CSF biomarkers as compared with traffic-related PM_2.5_ exposure. Ambient PM_2.5_ contains emissions from traffic, industry, domestic fuel burning, natural sources including soil dust and sea salt, as well as unspecific sources of human origin.^37^ On the other hand, traffic-related PM_2.5_ is a source of ambient PM_2.5_ that includes emissions of organic and inorganic gaseous PM precursors from the combustion of fuels and lubricants.^37^ Since both sources contain organic and often toxic particles, we expected to see relationships between both sources of PM_2.5_ and AD CSF biomarkers, and while not significant for Aβ_42_, the associations between traffic-related PM_2.5_ and AD CSF biomarkers were similar to associations with ambient PM_2.5_. More research needs to be done to determine which PM_2.5_ mixtures are particularly harmful to the central nervous system.

While we did not find effect modifications by *APOE*-ε4 carriership or other common risk factors for AD, the association between ambient PM_2.5_ exposure and Aβ_42_ CSF became stronger with increasing age (though not statistically significant). These results could suggest that AD CSF biomarkers might not be sensitive enough to detect AD-related changes in participants < 60 years old, but more research in the population will clarify the most clinically relevant age for biomarker measurement. Previous research suggests that biomarker patterns of Aβ_42_ consistent with stage 1 AD (amyloid pathology only) are first detectable during early middle age (45-54 years), while increases in tTau and pTau are typically not apparent until later (ages ≥ 55 years).^38^ However, this previous study used an unstandardized assay, the INNOTEST ELISA, which often yields systematic variability in comparison to the Elecsys assay. Another potential explanation for the stronger associations among participants older than 60 years could be the higher accumulative PM_2.5_ exposure over the lifetime among older individuals. In line with this hypothesis, one study examining the relationship between PM_2.5_ exposure and AD prevalence found a stronger effect of PM_2.5_ on AD prevalence among those at or above 70 years of age.^3^

There are several strengths to be noted, such as, the exposure assessment which included two sources of PM_2.5_, ambient and traffic-related, which were estimated at a high spatial resolution of up to 200m; our outcome assessment which relied on a recommended assay for AD CSF biomarker measurement,^16^ and for which we observed consistent associations using continuous biomarker concentrations as well as AD positivity cut-offs; our inclusion of several well-known confounders and methods to reduce confounding by neighborhood-level characteristics; and our relatively large sample size (N=1,113) of CSF measurements from cognitively healthy older adults free of chronic illness.

In addition to its strengths, this study has several limitations. Given that ADRD progresses over the course of several years or decades, we evaluated the associations with 3- and 5-year average PM_2.5_ concentrations prior to enrollment in addition to the 1-year averages. However, given that exposure was assigned based on the baseline residence and some participants could have relocated in the years prior to the study, the 3- and 5-year estimates may be affected by exposure misclassification. Such exposure misclassification is also a potential explanation for the weaker associations between the 3- and 5-year PM_2.5_ exposures and Aβ_42_ CSF concentrations in comparison to the 1-year exposure concentrations. Our study also only used cross-sectional CSF measurements; longitudinal repeated measures analyses may provide a better understanding of the long-term effect of air pollution on CSF biomarker trajectories of AD. Further, our sample was not representative of the Atlanta metropolitan area, the target population, as it was mainly high SES and white, which limits both the generalizability and transportability of our estimates. Finally, while we looked at two different sources of PM_2.5_, we did not examine the relationship between AD pathology and specific components of PM_2.5_. Future studies should consider the components of PM_2.5_ as they are dynamic between ambient and traffic-related sources with different toxicity^39^ and could reveal important and undiscovered relationships between exposure and disease pathogenesis.

In conclusion, our results suggest that exposure to ambient and traffic-related PM_2.5_, even at levels below current primary and secondary standards defined by the Environmental Protection Agency (EPA) for PM_2.5_ (annual average standards with levels of 12.0 μg/m^3^ and 15.0 μg/m^3^, respectively), increases the risk of future AD development. Additionally, our results add to the growing body of evidence which suggests that air pollution directly contributes to neurodegeneration by accelerating Aβ_42_ accumulation in the brain.^2, 40^

## Data Availability

All data produced in the present study are available upon reasonable request to the authors.

## Acknowledgments

We would like to thank Liuhua Shi and her team who provided us with guidance for the air pollution assignments of ambient PM2.5. We also want to thank Dr. Jeremy A. Sarnat (Emory), Dr. Armistead Russell (Georgia Tech), Ms. Kyung-Hwa Kim, and Ms. Abby Marinelli from the Atlanta Regional Commission for providing data and guidance towards the traffic-related air pollution exposure assessment.

