## Supplemental Material for "Association between Fine Particulate Matter Exposure and Cerebrospinal Fluid Biomarkers of Alzheimer’s Disease among a Cognitively Healthy Population-based Cohort"

**Table S1** Main results on the associations between exposure to PM_2.5_ and concentrations of Alzheimer’s disease (AD) Cerebrospinal fluid (CSF) biomarkers

| **Air Pollutant** | **AD CSF Biomarker** | **N** | **Beta** | **95% CI** | ***P*-value** |
| --- | --- | --- | --- | --- | --- |
| **1-year Ambient PM_2.5_** | **Aβ_42_** | **1113** | **-0.09** | **(-0.15; -0.02)** | **0.01** |
|  | **pTau** | 1113 | -0.05 | (-0.12; 0.01) | 0.11 |
|  | **tTau** | 1113 | -0.05 | (-0.12; 0.01) | 0.11 |
|  | **pTau/Aβ_42_** | 1113 | 0.05 | (-0.02; 0.11) | 0.18 |
|  | **tTau/Aβ_42_** | 1113 | 0.05 | (-0.02; 0.11) | 0.16 |
| **3-year Ambient PM_2.5_** | **Aβ_42_** | **1113** | **-0.07** | **(-0.13; -0.01)** | **0.03** |
|  | **pTau** | 1113 | -0.06 | (-0.11; 0.003) | 0.06 |
|  | **tTau** | **1113** | **-0.06** | **(-0.12; 0.001)** | **0.047** |
|  | **pTau/Aβ_42_** | 1113 | 0.02 | (-0.05; 0.08) | 0.62 |
|  | **tTau/Aβ_42_** | 1113 | 0.02 | (-0.04; 0.08) | 0.59 |
| **5-year Ambient PM_2.5_** | **Aβ_42_** | 1113 | -0.07 | (-0.13; 0.004) | 0.07 |
|  | **pTau** | 1113 | -0.05 | (-0.12; 0.02) | 0.14 |
|  | **tTau** | 1113 | -0.06 | (-0.12; 0.01) | 0.10 |
|  | **pTau/Aβ_42_** | 1113 | 0.02 | (-0.05; 0.09) | 0.57 |
|  | **tTau/Aβ_42_** | 1113 | 0.02 | (-0.05; 0.09) | 0.55 |
| **1-year Traffic-related PM_2.5_** | **Aβ_42_** | 1080 | -0.03 | (-0.10; 0.03) | 0.33 |
|  | **pTau** | 1080 | 0.01 | (-0.06; 0.8) | 0.77 |
|  | **tTau** | 1080 | 0.01 | (-0.06; 0.07) | 0.86 |
|  | **pTau/Aβ_42_** | 1080 | 0.04 | (-0.03; 0.11) | 0.25 |
|  | **tTau/Aβ_42_** | 1080 | 0.05 | (-0.02; 0.11) | 0.20 |
| **3-year Traffic-related PM_2.5_** | **Aβ_42_** | 1080 | -0.06 | (-0.13; 0.01) | 0.12 |
|  | **pTau** | 1080 | 0.002 | (-0.07; 0.07) | 0.96 |
|  | **tTau** | 1080 | -0.004 | (-0.07; 0.06) | 0.91 |
|  | **pTau/Aβ_42_** | 1080 | 0.05 | (-0.02; 0.12) | 0.14 |
|  | **tTau/Aβ_42_** | 1080 | 0.06 | (-0.01; 0.13) | 0.11 |
| **5-year Traffic-related PM_2.5_** | **Aβ_42_** | 1080 | -0.06 | (-0.14; 0.01) | 0.10 |
|  | **pTau** | 1080 | 0.01 | (-0.07; 0.08) | 0.90 |
|  | **tTau** | 1080 | -0.0003 | (-0.07; 0.07) | 0.99 |
|  | **pTau/Aβ_42_** | 1080 | 0.06 | (-0.01; 0.14) | 0.09 |
|  | **tTau/Aβ_42_** | 1080 | 0.07 | (-0.01; 0.14) | 0.07 |
| Associations were adjusted by gender, age, educational attainment, ethnic group, BMI, and neighborhood SES. Standardized coefficients are reported. Significant associations (p<0.05) are indicated in bold. | | | | | |

**Table S2** Main results on the associations between exposure to PM_2.5_ and AD positive (+) cut-offs for AD CSF biomarkers

| **Air Pollutant** | **AD CSF (+) cut-offs** | **N** | **Odds Ratio** | **95% CI** | ***P*-value** |
| --- | --- | --- | --- | --- | --- |
| **1-year Ambient PM_2.5_** | **Aβ_42_** | **1113** | **1.23** | **(1.07; 1.42)** | **0.004** |
|  | **pTau** | 1113 | 0.11 | (0.82; 1.42) | 0.59 |
|  | **tTau** | 1113 | 0.92 | (0.70; 1.20) | 0.52 |
|  | **pTau/Aβ_42_** | 1113 | 1.15 | (0.93; 1.43) | 0.20 |
|  | **tTau/Aβ_42_** | 1113 | 1.20 | (0.94; 1.55) | 0.13 |
| **3-year Ambient PM_2.5_** | **Aβ_42_** | **1113** | **1.20** | **(1.04; 1.37)** | **0.009** |
|  | **pTau** | 1113 | 1.01 | (0.78;1.39) | 0.65 |
|  | **tTau** | 1113 | 0.92 | (0.71; 1.17) | 0.48 |
|  | **pTau/Aβ_42_** | 1113 | 1.09 | (0.89; 1.33) | 0.43 |
|  | **tTau/Aβ_42_** | 1113 | 1.13 | (0.89; 1.42) | 0.31 |
| **5-year Ambient PM_2.5_** | **Aβ_42_** | **1113** | **1.21** | **(1.04; 1.40)** | **0.01** |
|  | **pTau** | 1113 | 1.07 | (0.80,1.43) | 0.65 |
|  | **tTau** | 1113 | 0.95 | (0.71; 1.26) | 0.71 |
|  | **pTau/Aβ_42_** | 1113 | 1.11 | (0.89; 1.40) | 0.36 |
|  | **tTau/Aβ_42_** | 1113 | 1.21 | (0.93; 1.58) | 0.16 |
| **1-year Traffic-related PM_2.5_** | **Aβ_42_** | 1080 | 1.10 | (0.95; 1.27) | 0.22 |
|  | **pTau** | 1080 | 0.97 | (0.58; 1.60) | 0.89 |
|  | **tTau** | 1080 | 1.02 | (0.75; 1.38) | 0.89 |
|  | **pTau/Aβ_42_** | 1080 | 1.35 | (0.94; 1.94) | 0.10 |
|  | **tTau/Aβ_42_** | 1080 | 1.26 | (0.84; 1.90) | 0.27 |
| **3-year Traffic-related PM_2.5_** | **Aβ_42_** | 1080 | 1.14 | (0.98; 1.32) | 0.10 |
|  | **pTau** | 1080 | 0.95 | (0.56; 1.62) | 0.85 |
|  | **tTau** | 1080 | 0.98 | (0.71; 1.34) | 0.88 |
|  | **pTau/Aβ_42_** | **1080** | **1.52** | **(1.05; 2.20)** | **0.03** |
|  | **tTau/Aβ_42_** | 1080 | 1.41 | (0.93; 2.15) | 0.11 |
| **5-year Traffic-related PM_2.5_** | **Aβ_42_** | 1080 | 1.15 | (0.98; 1.35) | 0.08 |
|  | **pTau** | 1080 | 0.98 | (0.56; 1.70) | 0.95 |
|  | **tTau** | 1080 | 0.10 | (0.71; 1.40) | 0.10 |
|  | **pTau/Aβ_42_** | **1080** | **1.56** | **(1.06; 2.28)** | **0.02** |
|  | **tTau/Aβ_42_** | 1080 | 1.44 | (0.93; 2.22) | 0.10 |
| Associations were adjusted by gender, age, educational attainment, ethnic group, BMI, and neighborhood SES. Significant associations (p<0.05) are indicated in bold. **Table S3** Results on the associations between exposure to Ambient PM_2.5_ and concentrations of Alzheimer’s disease (AD) Cerebrospinal fluid (CSF) biomarkers (linear: continuous biomarker concentrations; logistic: AD CSF cut-offs) for participants with traffic-related exposure, located in the metro-Atlanta area (N=1,080) **Air Pollutant** \| **AD CSF Biomarker** \| **N** \| **Beta** \| **95% CI** \| ***P*-value**; **1-year Ambient PM_2.5_** \| **Aβ_42_** \| 1080 \| -0.09 \| (-0.16; -0.02) \| 0.01; \| **pTau** \| 1080 \| -0.06 \| (-0.13; 0.01) \| 0.10; \| **tTau** \| 1080 \| -0.06 \| (-0.12; 0.01) \| 0.10; \| **pTau/Aβ_42_** \| 1080 \| 0.05 \| (-0.02; 0.12) \| 0.17; \| **tTau/Aβ_42_** \| 1080 \| 0.05 \| (-0.02; 0.12) \| 0.16; **3-year Ambient PM_2.5_** \| **Aβ_42_** \| 1080 \| -0.07 \| (-0.13; -0.004) \| 0.04; \| **pTau** \| 1080 \| -0.07 \| (-0.13; -0.003) \| 0.04; \| **tTau** \| 1080 \| -0.07 \| (-0.13; -0.01) \| 0.03; \| **pTau/Aβ_42_** \| 1080 \| 0.05 \| (-0.02; 0.11) \| 0.12; \| **tTau/Aβ_42_** \| 1080 \| 0.01 \| (-0.05; 0.08) \| 0.67; **5-year Ambient PM_2.5_** \| **Aβ_42_** \| 1080 \| -0.07 \| (-0.14; -0.01) \| 0.08; \| **pTau** \| 1080 \| -0.06 \| (-0.13; 0.01) \| 0.10; \| **tTau** \| 1080 \| -0.07 \| (-0.14; 0.01) \| 0.07; \| **pTau/Aβ_42_** \| 1080 \| 0.02 \| (-0.06; 0.09) \| 0.66; \| **tTau/Aβ_42_** \| 1080 \| 0.02 \| (-0.06; 0.09) \| 0.64; **Air Pollutant** \| **AD CSF Biomarker** \| **N** \| **Odds Ratio** \| **95% CI** \| ***P*-value**; **1-year Ambient PM_2.5_** \| **Aβ_42_** \| 1080 \| 1.24 \| (1.07; 1.44) \| 0.005; \| **pTau** \| 1080 \| 1.02 \| (0.76; 1.38) \| 0.88; \| **tTau** \| 1080 \| 0.80 \| (0.60; 1.07) \| 0.13; \| **pTau/Aβ_42_** \| 1080 \| 1.20 \| (0.95; 1.51) \| 0.12; \| **tTau/Aβ_42_** \| 1080 \| 1.28 \| (0.98; 1.68) \| 0.07; **3-year Ambient PM_2.5_** \| **Aβ_42_** \| 1080 \| 1.20 \| (1.05; 1.39) \| 0.01; \| **pTau** \| 1080 \| 0.94 \| (0.71; 1.23) \| 0.63; \| **tTau** \| 1080 \| 0.81 \| (0.62; 1.06) \| 0.12; \| **pTau/Aβ_42_** \| 1080 \| 1.12 \| (0.90; 1.40) \| 0.30; \| **tTau/Aβ_42_** \| 1080 \| 1.18 \| (0.91; 1.53) \| 0.21; **5-year Ambient PM_2.5_** \| **Aβ_42_** \| 1080 \| 1.20 \| (1.03; 1.41) \| 0.02; \| **pTau** \| 1080 \| 0.99 \| (0.73; 1.36) \| 0.97; \| **tTau** \| 1080 \| 0.82 \| (0.61; 1.11) \| 0.21; \| **pTau/Aβ_42_** \| 1080 \| 1.15 \| (0.90; 1.48) \| 0.26; \| **tTau/Aβ_42_** \| 1080 \| 1.27 \| (0.95; 1.70) \| 0.10; Associations were adjusted by gender, age, educational attainment, ethnic group, BMI, and neighborhood SES. Standardized coefficients are reported. Significant associations (p<0.05) are indicated in bold. \| \| \| \| \| **Table S4** Results on the associations between annual exposure to Ambient PM_2.5_ and concentrations of Alzheimer’s disease (AD) Cerebrospinal fluid (CSF) biomarkers by common risk factors **Covariate** \| **1-year Ambient PM_2.5_** \| **N** \| **Interaction P-value**; **APOE-e4 status** \| \| \| 0.59; Yes \| -0.16 (-0.30; -0.02) \| 264 \|; No \| -0.11 (-0.22; -0.01) \| 591 \|; Overall \| -0.13 (-0.21;-0.04) \| 855 \|; **Family History of AD** \| \| \| 0.37; Yes \| -0.16 (-0.32; -0.01) \| 252 \|; No \| -0.09 (-0.18; 0.004) \| 861 \|; Overall \| -0.10 (-0.18; -0.03) \| 1113 \|; **ADI** ≥50 <50 Overall \| \| \| 0.62; \| -0.06 (-0.24, 0.12) \| 184 \|; \| -0.11 (-0.20; -0.03) \| 929 \|; \| -0.10 (-0.18; -0.02) \| 1113 \|; **Gender** Male \| \| \| 0.67; \| -0.13 (-0.26; 0.01) \| 338 \|; Female \| -0.09 (-0.18; 0.004) \| 775 \|; Overall \| -0.10 (-0.18; -0.02) \| 1113 \| **Table S5** Results (Odds Ratio ± 95% CI) on the associations between annual exposure to Ambient PM_2.5_ and concentrations of Alzheimer’s disease (AD) Cerebrospinal fluid (CSF) biomarkers by common risk factors **Covariate** \| **1-year Ambient PM_2.5_** \| **N** \| \| **Interaction P-value**; **APOE-e4 status** \| \| \| 0.80 \|; Yes \| 1.48 (1.08; 2.02) \| 264 \| \|; No \| 1.40 (1.08; 1.80) \| 591 \| \|; Overall \| 1.43 (1.17; 1.74) \| 855 \| \|; **Family History of AD** \| \| \| \| 0.27; Yes \| 1.51 (1.08; 2.09) \| 252 \| \|; No \| 1.21 (1.00; 1.48) \| 861 \| \|; Overall \| 1.29 (1.09; 1.53) \| 1113 \| \|; **ADI** ≥50 <50 Overall \| \| \| \| 0.12; \| 0.96 (0.64; 1.43) \| 184 \| \|; \| 1.36 (1.13; 1.64) \| 929 \| \|; \| 1.28 (1.08; 1.52) \| 1113 \| \|; **Gender** Male \| \| \| \| 0.72; \| 1.34 (1.00; 1.79) \| 338 \| \|; Female \| 1.25 (1.02; 1.79) \| 775 \| \|; Overall \| 1.28 (1.08; 1.52) \| 1113 \| \| **Table S6** Neighborhood-level characteristics used in the construction of neighborhood deprivation principal components **Potential confounder Neighborhood-level Characteristics** \| Overall (N=1113); **% Education < High School** \|; Mean (SD) \| 8.07 (7.23); **Unemployment Rate** \|; Mean (SD) \| 7.38 (4.50); **% Males not in Labor Force** \|; Mean (SD) \| 24.8 (8.17); **% Vacant Housing** \|; Mean (SD) \| 9.22 (5.96); **% Renter Occupied** \|; Mean (SD) \| 31.9 (20.8); **% Crowded Household** \|; Mean (SD) \| 1.62 (2.15); **Median Household Value ($)** \|; Mean (SD) \| 268000 (154000); **% Males not in management, etc.** \|; Mean (SD) \| 52.7 (18.8); **% Females not in management, etc.** \|; Mean (SD) \| 48.7 (14.7); **% Individuals in Poverty** \|; Mean (SD) \| 8.53 (7.84); **% Female Headed Household** \|; Mean (SD) \| 6.17 (4.59); **% Household < $35,000 annual income** \|; Mean (SD) \| 24.0 (12.4); **% Household on public assistance** \|; Mean (SD) \| 1.31 (1.45); **% Household without vehicles** \|; Mean (SD) \| 13.7 (15.9); **% Non-Hispanic Black** \|; Mean (SD) \| 22.9 (24.4); **% Hispanic or Latino Origin** \|; Mean (SD) \| 8.51 (9.41) | | | | | |

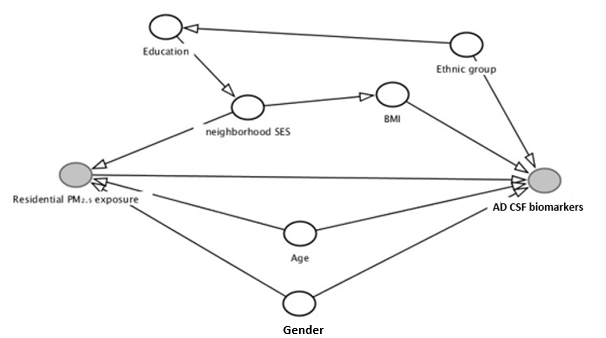

**Figure S1** Directed Acyclic Graph (DAG) connecting PM_2.5_ to AD CSF biomarkers and defining potential confounding factors. PM_2.5_, fine particulate matter; AD; Alzheimer’s disease; CSF, cerebrospinal fluid; BMI, body mass index; SES, socioeconomic status

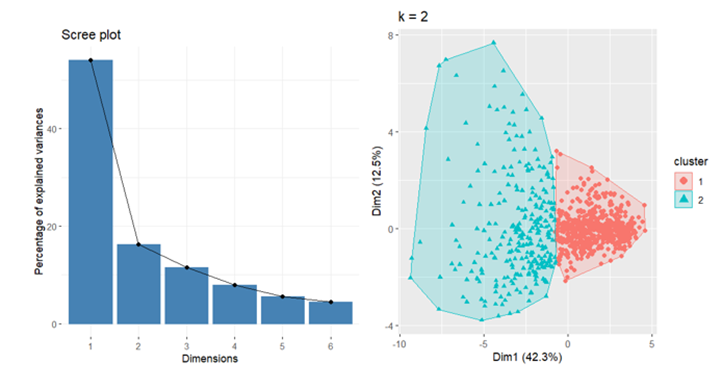

**Figure S2** Elbow method and results for computing neighborhood deprivation indicators.

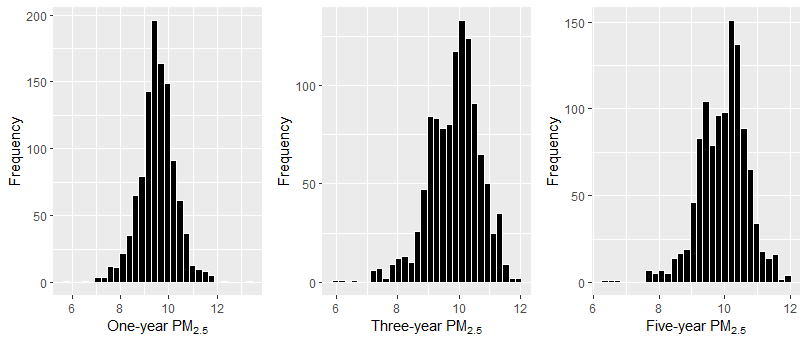

**Figure S3** Histograms of 1, 3, and 5-year ambient PM_2.5_ exposure concentrations (in μg/m^3^).

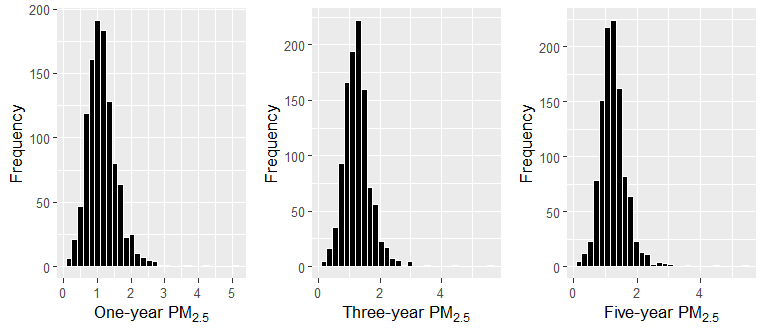

**Figure S4** Histograms of 1, 3, and 5-year traffic-related PM_2.5_ exposure concentrations (in μg/m^3^).

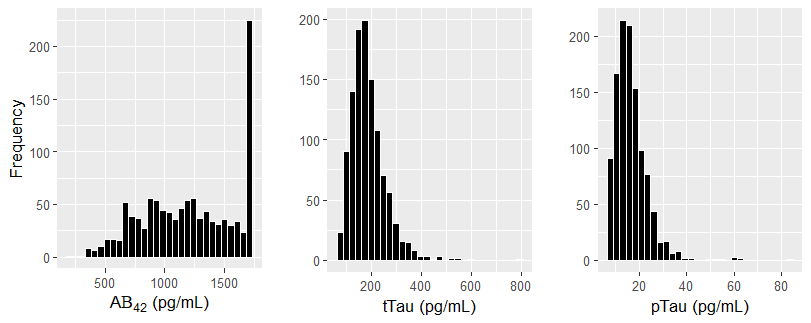

**Figure S5** Histograms of Aβ_42_, tTau, and pTau concentrations (in pg/mL) collected at baseline (2016-2020) before log transformation.

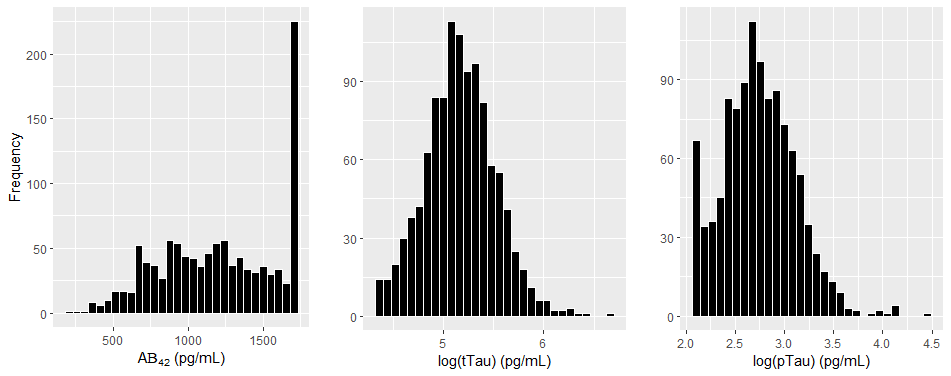

**Figure S6** Histograms of Aβ_42_, tTau, and pTau concentrations (in pg/mL) collected at baseline (2016-2020) after log transformation.

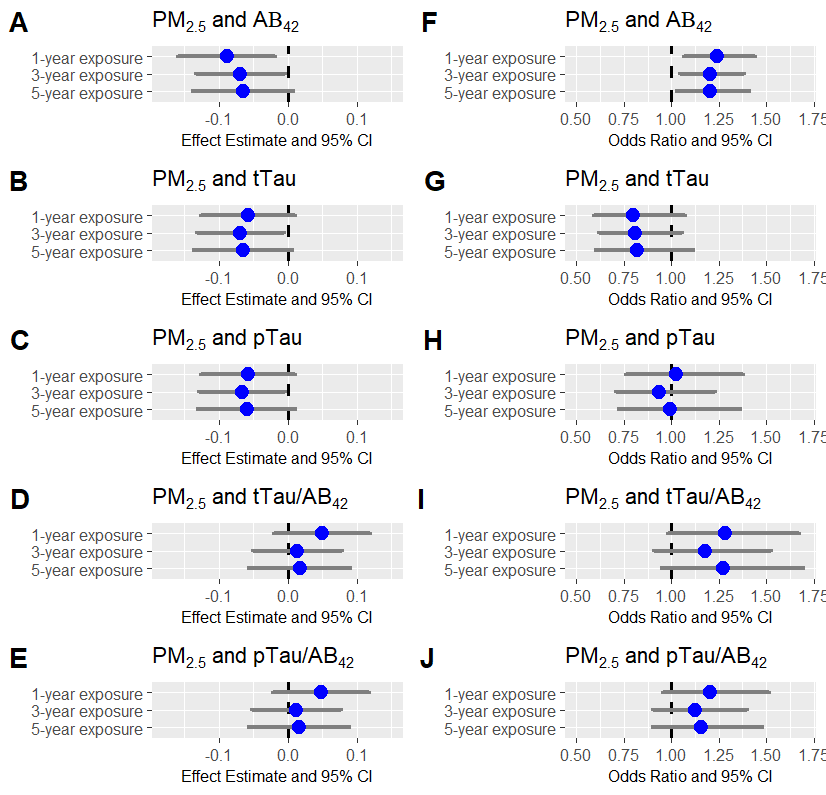

**Figure S7** Effect Estimate (± 95% CI) **A**–**E** and Odds ratio (± 95% CI) **F**–**J** for the association between 1, 3, and 5-year ambient PM_2.5_ exposure and AD CSF Biomarker (Aβ_42_, tTau, pTau, tTau/Aβ_42_, and pTau/Aβ_42_) concentrations or AD positive (+) cut-offs. Sample size (**N=1080**) represents the subset of EHBS participants with available traffic-related PM_2.5_ exposure at the residence. All estimates are standardized and adjusted for gender, age, N-SES, ethnicity, educational attainment, and BMI. The dashed line indicates the significance threshold: 0 for linear regression and 1 for logistic regression.

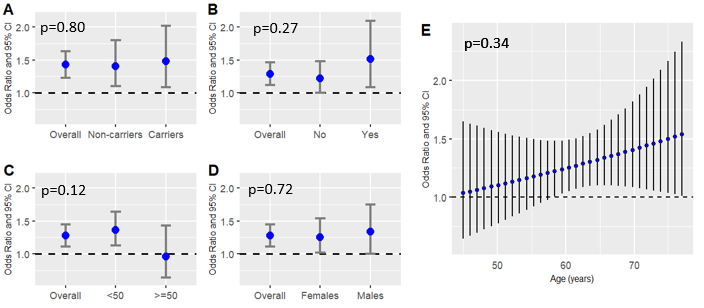

**Figure S8** Odds Ratio (± 95% CI) for the association between yearly ambient PM_2.5_ exposure and Aβ_42_ AD CSF Biomarkers positive (+) cut-offs by *APOE*-ε4 carriership (**A**), AD family history (**B**), ADI (**C**), Gender (**D**), and Age (**E**). Presented as overall and stratified effects for dichotomous variables and as continuous for Age, with interaction p-values depicted on each graph. The dashed line indicates the significance threshold: 1 for logistic regression. The overall effect in **Figure S8A** (N=855) differs slightly from **Figure S8B-D** (N=1113) due to the decreased sample size after including only participants with *APOE* genotype data.
